# Assessing the Impact of use of HIV self-testing on the incidence of HIV Infection in Nigeria: a systematic review and meta-analysis

**DOI:** 10.1101/2024.04.04.24305344

**Authors:** George Uchenna Eleje, Godwin Omokhagbo Emmanuel, Folahanmi Tomiwa Akinsolu, Morẹ́nikẹ́ Oluwátóyìn Foláyan

## Abstract

**Background:** Human immunodeficiency virus (HIV) self-testing tool is a widely adopted tool in Nigeria. However, there is little known about its impact in reducing HIV infection rates in Nigeria. This review aims to assess the impact of the use of HIV self-testing on the incidence of HIV infections in the country.

**Methods:** This was a systematic review and meta-analysis. Studies conducted in Nigeria on HIV self-testing with or without comparison to other HIV tests were included. The primary outcomes considered were the detection rate of new HIV cases and the acceptability (uptake) rate for HIV self-testing. Secondary outcomes were the usability rate, repeat testing rate, willingness rate, awareness rate, incidence of social harm, and incidence of high-risk behaviour. Electronic databases (PubMed/Medline, Web of Science, Scopus, CINAHL, and Cochrane Library) and Google Scholar were searched for relevant studies. Searches were conducted till December 2, 2023. Pooled estimates were calculated using a random-effects model with the DerSimonian Laird method. Heterogeneity was analyzed using the I^2^ test, and risk of bias was assessed with the Hoy and colleagues’ scale. Meta-analysis was conducted where possible. The protocol was registered with PROSPERO (CRD42023479752).

**Results:** Eight studies, encompassing 7,556 participants, met the inclusion criteria. The overall risk of bias for the included studies was adjudged low. The detection rate of HIV self-testing for new HIV cases was 25.78% (95% CI: 0.90-50.66, I^2^:100.0), acceptability (uptake) rate was 56.92% (95% CI: 26.54-87.30, I^2^:100.0), and repeat testing rate was 20.10% (95% CI: -11.44-51.65, I^2^:100.0). Usability rate, willingness rate, awareness rate, and incidence of high-risk behaviour were reported in one study respectively, with no information on the incidence of social harm. Sensitivity analysis was done, and subgroup analyses could not be estimated due to insufficient data.

**Conclusions:** The use of HIV self-testing test kits in Nigeria showed a high detection rate of new HIV cases, moderate acceptability, but low repeat testing rates. However, the evidence is limited. Larger, higher-quality studies are essential to explore the broader impact of HIV self-testing on reducing HIV incidence in Nigeria.

## Introduction

Nigeria, in 2018, had an estimated HIV incidence rate of 0.36 per 1,000 population for all age [1]. Of the 77,200 new infections estimated in Nigeria by the end of 2022, adult women (age 15 years and above) accounted for 35,000 (45.3%) [1]. Nigeria however, recorded a 39% reduction in new HIV infections between 2010 and 2022 [1]. Though the prevalence of HIV in Nigeria in 2018 was estimated as 1.3% among adults aged 15 to 49, the prevalence was 1.43% compared to 0.37% for young people aged 15-24 years. Among adults, females have a higher prevalence compared to males (1.75% vs. 0.95%) The HIV incidence-to-prevalence ratio in 2022 was 3.85, whereas the benchmark for epidemic control is a ratio of 3% [2,3]. More recent estimated for 2022 suggests that the incidence of HIV is increasing with a 2022 estimated national HIV prevalence of 2.1% (95% CI: 1.5–2.7%) among adults aged 15–49 years in Nigeria, which corresponds to approximately 2 million people living with HIV [4]. This suggests that Nigeria needs to considerably strengthen its HIV control effort before it can achieve “epidemic control.”

HIV/AIDS remains a public health concern in Nigeria. A critical point of strengthening the HIV response in Nigeria is through improved HIV testing. Rapid tests for recent infection identify people with new HIV infections, and the aggregated data can identify hot spots of current HIV transmission. A strategic approach for HIV epidemic control is promoting the uptake of HIV testing services [5]. Stigma and poor hospital access has limited uptake of conventional HIV services [6]. The use of self-test kits helps overcome some of these barriers and may have contributed significantly to HIV control in Nigeria. The magnitude of this contribution remains unknown.

HIV self-testing had been the main HIV self-test kit in Nigeria since 2019 with over 3,000,000 kits distributed in Nigeria since introduction. HIV self-test was approved by the FDA in 2002 and WHO in 2017 for use in detecting antibodies to the Human Immunodeficiency Virus Type 2, or HIV-2, in oral fluid samples in the US and globally respectively. Similar approval was gotten from NAFDAC in 2019 for use in Nigeria. Its global proven and documented performance is a sensitivity 99.4%, specificity 99.0%, usability score of 98% and detection of IGG/IgM antibodies 25 days from infection and can be compared with 4th Generation EIA.

The WHO recommends HIV self-testing as an innovative strategy and an additional testing approach to attain UNAIDS targets to end HIV by 2030 [7].

In the quest to reduce HIV incidence, the introduction of novel HIV self-testing kits is of growing interest. The evidence gap of HIV self-testing kit could be partly addressed by synthesizing and pooling estimates from existing country-level evidence via systematic review methods and meta-analysis. The aim of this study is to map the use of HIV self-testing for HIV prevention programming in Nigeria, and to estimate the impact of the introduction of self-test kits in promoting access to HIV testing and new case finding in Nigeria.

## Methods

### Study protocol

The research adhered to a pre-established protocol for systematic review and meta-analysis. Registration was completed with PROSPERO under the number: CRD42023479752. The study was documented in accordance with the Preferred Reporting Items for Systematic Reviews and Meta-Analyses (PRISMA) statement and checklist. Two blinded authors independently conducted each stage of the review, resolving disagreements through discussion with a third author.

### Search strategy

Searches were conducted from the inception of six electronic databases spanning from their inception to December 2023. Those on electronic databases were PubMed/Medline, Web of Science, Scopus, CINAHL, and the Cochrane Library and Google Scholar. The initial search syntax was initially formulated for PubMed and subsequently adjusted to meet the distinct search criteria of the other databases. The search strategies can be found in Appendix 1. The review process entailed the assessment of abstracts from all the references extracted from eligible articles. Additionally, supplementary articles were identified by scrutinizing the reference lists of already recognized articles.

### Study selection

Two researchers (GUE and FTA) independently assessed and identified the studies for inclusion in the review, considering the predetermined inclusion and exclusion criteria. The titles and abstracts of all studies underwent initial screening, followed by a comprehensive evaluation of the full texts of the selected studies to determine eligibility. A comparison of independently selected articles by the two authors was conducted. Discrepancies were addressed through a collaborative decision-making process in consultation with the third author, MOF, to resolve disagreements concerning the selected articles.

### Eligibility criteria

The systematic review and meta-analysis employed the Condition, Context, and Population (CoCoPop) method for formulating research questions [8]. Inclusion criteria comprised studies conducted in Nigeria that assessed the magnitude and/or determinants of HIV self-testing kits.

The study design had to be observational (cross-sectional, case-control, or cohort studies), and the selected studies needed to report on at least one of the following outcomes: detection rate of new HIV cases, acceptability (uptake) rate, usability rate, repeat testing rate, willingness rate, awareness rate, incidence of social harm, or incidence of high-risk behavior.

The review was limited to quantitative, peer-reviewed studies - whether published or unpublished – studies. In instances of duplicate studies derived from the same dataset, the most comprehensive and current version was included. Eligible studies fell within the categories of cross-sectional studies, cohort studies, case-control studies, or randomized controlled or controlled clinical studies. Additionally, studies were included if they presented available data for at least one of the primary outcomes. There was no language restriction. Studies excluded were case reports, case series, commentaries, conference abstracts, letters to editors, technical reports, review articles, qualitative studies, and other opinion publications. Studies lacking details on sample size, those conducted outside Nigeria, studies with inaccurate or unavailable outcome data, and studies featuring duplicate samples were also excluded.

### Quality and risk of bias within studies assessment

Risk of bias assessments were conducted using Cochrane’s risk of bias tool version 2.0 (RoB 2.0) for randomized control trials [9] and the risk of bias tool developed by Hoy and colleagues scale for other study designs [10]. Only articles deemed to have low or moderate risk of bias were incorporated into the meta-analysis. Two authors independently assessed each study using the critical appraisal checklist, with the third author verifying the evaluations.

### Outcome measurements

The primary outcome measures were: (1) the detection rate of new cases of HIV infections using HIV self-testing in Nigeria; and (2) the acceptability (uptake) rate of HIV self-testing in Nigeria. The secondary outcome measures were (1) the usability rate of HIV self-testing in Nigeria; (2) the repeat testing rate of using HIV self-testing methods in Nigeria; (3) the incidence of social harm using HIV self-tests methods in Nigeria; and the (4) incidence of high-risk behaviour following HIV self-testing methods in Nigeria.

### Subgroup and sensitivity analyses

Subgroup analyses were planned to identify potential sources of heterogeneity. Planned subgroups were based on age (children and adults or adolescents and adults), sex work (sex workers and non-sex workers) and sex (males and females). A subgroup effect was deemed present when the interaction test in Review Manager 5.4.1 (The Nordic Cochrane Center, Copenhagen, Denmark) indicated significant group differences (p < 0.10). A leave-out-one sensitivity analyses were performed to explore the impact of each study on the pooled results from the meta-analysis while gradually excluding each study.

### Assessment for publication bias

Publication bias and the potential impact of small studies were planned to be evaluated employing funnel plots and Egger’s tests for outcomes encompassing at least ten studies.

### Data extraction

A standardized data extraction form was developed to gather information from each study included in the systematic review. The data extracted included details on the name of the first author of the publication, year of publication, the year of data collection, the region where the study was conducted in Nigeria (Northern or Southern), the city of the study, the study setting, the study design, the count of cases, the mean or median age of the participants, uptake rates, sample size, tools used for data collection, sex distribution among cases, and the study population. In cases where information was missing, the authors endeavoured to obtain this directly from the original authors via email when feasible.

### Data synthesis and analysis

The analysis was conducted using Review Manager 5.4.1 (Copenhagen: The Nordic Cochrane Centre, Cochrane Collaboration, 2021). These uptake rate values were visually presented on a forest plot. The DerSimonian-Laird random effects model was employed across all meta-analyses. We pooled data about the reported outcome rates and percentage (with 95% confidence interval) was used as the effect size, and then the inverse variance method (Generic Inverse Variance) was selected to calculate the pooled effect. Because, in this statistical procedure, the rates difference (RD) and its standard error are noted to be equivalent to the effect of a single rate and the standard error. To assess study heterogeneity, the Cochran’s (Q) statistic test, I^2^ statistic (Higgins and Thompson’s I^2^ statistics) and forest plots were utilized. The extent of statistical heterogeneity among studies was assessed, with categorizations of null (I^2^ = 0), insignificant (0 < I^2^ ≤ 25%), low (25 < I^2^ ≤ 50%), moderate (50 < I^2^ ≤ 75%), and high (I^2^ > 75%) heterogeneity.

## Results

### Study selection

The PRISMA flow chart outlining the results of the literature searches is shown in Figure 1.

**Figure 1:**
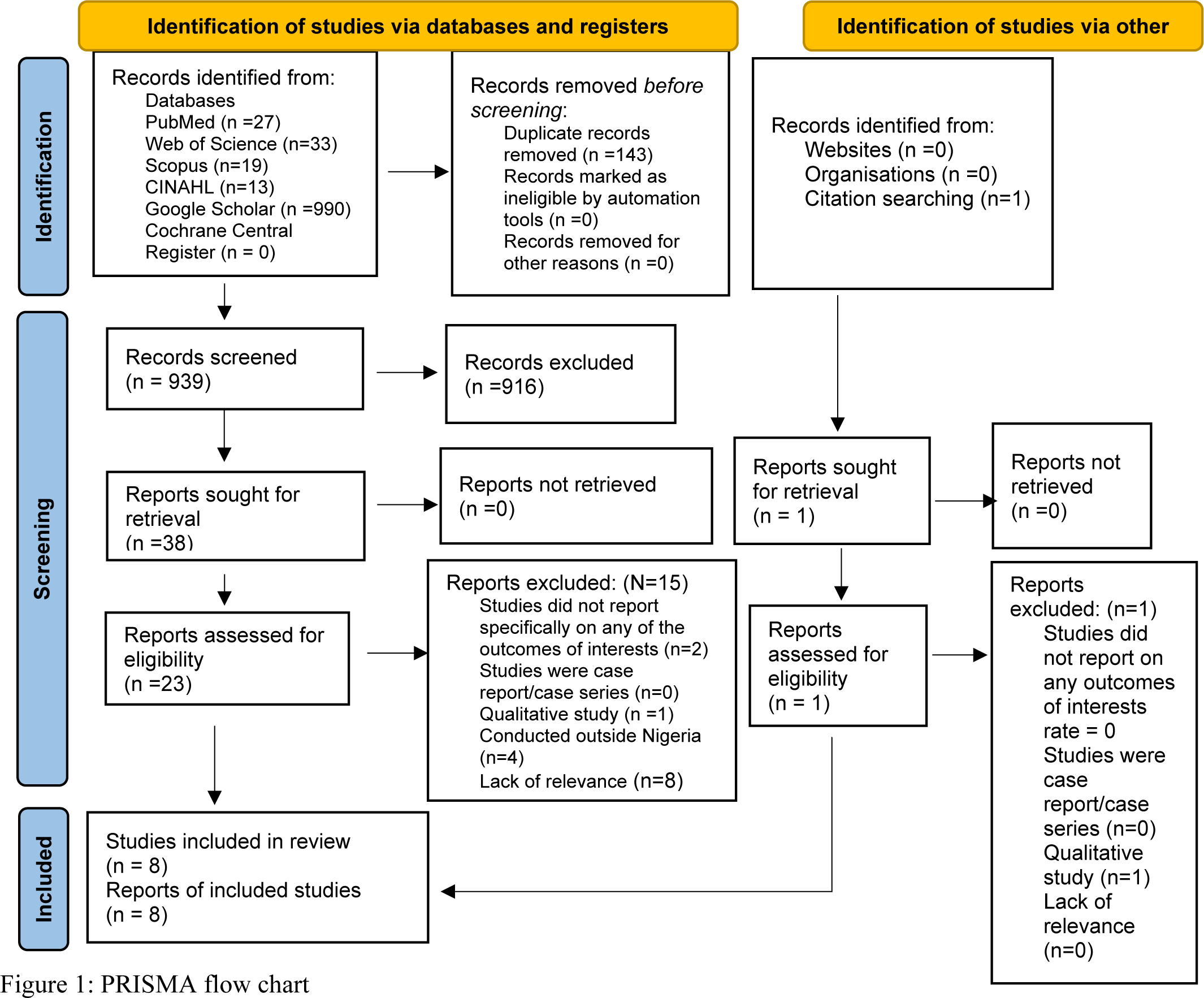
PRISMA flow chart

Twenty-four full-text studies were evaluated for eligibility, all of which were published in English. No gray literature or unpublished manuscripts were identified. As indicated in Table 1, eight studies met the criteria for inclusion [11–18]. Each of these included studies focused on oral HIV self-testing, with two specifically reporting on OraQuick [14, 17]. Conversely, sixteen papers were excluded, as illustrated in Table 2 [19–34].

**TABLE 1.** Summary of the characteristics of the studies included in this review

| Study | Oraquick-specific | Comparison with other HIV test kit | Period of participants' recruitment | State (Region) of recruitment | Site of study | Population | Sample size (M vs F) | Study design | Mean or median age | Age range (years) |
| --- | --- | --- | --- | --- | --- | --- | --- | --- | --- | --- |
| Adebimpe et al (2022) [11] | No | No | NR | Delta State (Southern Nigeria) | Ekpan General Hospital | Women of reproductive age attending immunization clinic | 357 (All F) | Cross sectional | 33.6 ±7.3 years | 31-40 |
| Agada et al (2021) [12] | No | No | March to December 2020 | Cross River and Akwa Ibom States (Southern Nigeria) | Community pharmacy | Patients | 5153 (2809 vs 2344) | Retrospective cross sectional | NR | 20-39 |
| Babatunde et al (2022) [13] | No | No | September 2020 | Oyo State (Southern Nigeria) | UI and LAUTECH, Ogbomoso | Undergraduate students | 155 82 vs 72; one missing data) | Cross sectional | NR | 16-30 |
| Iliyasu et al (2020) [14] | No | No | NR | Kano State (Northern Nigeria) | Bayero University/AKTH | University students | 399 (239 vs 160) | Cross sectional | 23.3±0.18 years | 20-30 |
| Iwelunmor et al (2022) [15] | No | No | September 2019 to March 2020 | Lagos, Enugu, Ondo, Oyo States (Southern Nigeria) | NR | Youth | 388 (NR) | Quasi experimental | NR | 14-24 |
| Nwoga et al (2012) [16] | Yes | Yes | April 2002 to January 2003 | Lagos State (Southern Nigeria) | LUTH, Nigeria | Patients | 281 (119 vs 162) | Cross sectional | 34.8 years | 1.5 – 75 |
| Ong et al (2022) [17] | No | Yes | December 2018 to February 2019 | Lagos State (Southern Nigeria) | Technical colleges, institutional campuses, and open community settings | Youth | 504 (NR) | Discrete choice Experiment | 21.0 ±2.0 years | 14-24 |
| Tun et al (2018) [18] | Yes | No | May to September 2017 | Lagos State (Southern Nigeria) | Population Council's MSM-friendly community-based health center, Lagos | MSM | 319 (All M) | Cohort | 25 years | 14-24 |

Abbreviations: NR=Not reported; AKTH=Aminu Kano Teaching Hospital; LUTH=Lagos University Teaching Hospital, Nigeria; LAUTECH= Ladoke Akintola University of Technology; UI=University of Ibadan; MSM=men who have sex with men (MSM); M=Male; F=Female

**Table 2:** Reasons for Exclusion.

|  | **Author and Year** | **Study Title** | **Reason for Exclusion** |
| --- | --- | --- | --- |
| 1. | Adeoti, 2021 [19] | Sexual practices, risk perception and HIV self-testing acceptability among long-distance truck drivers in Ekiti State, Nigeria | Lack of relevance to the research question - do not directly address the hypothesis of the review. |
| 2. | Adepoju, 2022 [20] | How efficient are HIV self-testing models? A comparison of community, facility, one-stop-shop and pharmacy retail distribution models in Nigeria | Lack of relevance to the research question - do not directly address the hypothesis of the review. |
| 3. | Aizobu, 2023 [21] | Enablers and barriers to effective HIV self-testing in the private sector among sexually active youths in Nigeria: A qualitative study using journey map methodology | Qualitative Study |
| 4. | Brown, 2023 [22] | HIV self-testing in Nigeria: public opinions and perspectives. | Public opinion poll (Qualitative study) |
| 5. | Dirisu, 2020 [23] | I will welcome this one 101%, I will so embrace it': a qualitative exploration of the feasibility and acceptability of HIV self-testing among men who have sex with men (MSM) in Lagos, Nigeria. | Lack of relevance to the research question - do not directly address the hypothesis of the review. |
| 6. | Hensen, 2018 [24] | Community-based distribution of oral HIV self-testing kits | Conducted outside Nigeria |
| 7. | Mavhu, 2022 [25] | Preferences for oral-fluid-based or blood-based HIV self-testing and provider-delivered testing: an observational study among different populations in Zimbabwe | Conducted outside Nigeria |
| 8. | Negedu, 2023 [26] | HIV self-testing among young persons in Nigeria: Knowledge, perceptions and potential for use | Conference Abstract - provide preliminary findings and lack comprehensive data |
| 9. | Nwaozuru, 2020 [27] | An innovation bootcamp model to develop HIV self-testing social enterprise among young people in Nigeria: a youth participatory design approach | Lack of relevance to the research question - do not directly address the hypothesis of the review. |
| 10. | Nwaozuru, 2021 [28] | Tailoring youth-friendly health services in Nigeria: a mixed-methods analysis of a designathon approach. | Lack of relevance to the research question - do not directly address the hypothesis of the review. |
| 11. | Oladele, 2023 [29] | An Unstructured Supplementary Service Data System to Verify HIV Self-Testing Among Nigerian Youths: Mixed Methods Analysis of Usability and Feasibility | Lack of relevance to the research question - do not directly address the hypothesis of the review. |
| 12. | Rosenberg, 2021 [30] | Strategies for enhancing uptake of HIV self-testing among Nigerian youths: a descriptive analysis of the 4YouthByYouth crowdsourcing contest | Study Design - Do not meet specific design criteria set out for the review |
| 13. | Stacey, 2022 [31] | Understanding factors that promote uptake of HIV self-testing among young people in Nigeria: Framing youth narratives using the PEN-3 cultural model. | Lack of relevance to the research question - do not directly address the hypothesis of the review. |
| 14. | Tahlil, 2021 [32] | A designathon to co-create community-driven HIV self-testing services for Nigerian youth: findings from a participatory event. | Lack of relevance to the research question - do not directly address the hypothesis of the review. |
| 15. | Young, 2014 [33] | Acceptability of Using Electronic Vending Machines to Deliver Oral Rapid HIV Self-Testing | Conducted outside Nigeria |
| 16. | Zhou, 2023 [34] | The roles of two HIV self-testing models in promoting HIV-testing among men who have sex with men | Conducted outside Nigeria |

### Characteristics of included studies

Table 1 presents the details of the studies included into the systematic review. Six of the eight studies provided information on the period of participants’ recruitment, spanning from April 2002 to December 2020 [12, 13, 15–18]. The study designs included five cross-sectional studies [11–14, 16], one cohort study [18], and two quasi-experimental studies [15, 17]. Babatunde et al [12] conducted an online assessment. All studies contributed at least one relevant dataset for analysis.

The sample size ranged from 155 to 5153. Additionally, the study population ranged from patients [12, 16] to non-patient [11, 13, 14, 17, 18]; and from youth [15, 17] to exclusively female populations [11] or male populations [18] and tertiary institution student populations [13, 14]. Tun et al [18] exclusively included men who have sex with men (MSM). The studies collectively enrolled a total of 7,556 participants. This included 3568 males and 3095 females. The sex of 893 participants were not reported [15, 17].

Geographically, seven of the eight studies were conducted in southern Nigeria [11–13, 15–18], and one study was conducted in Northern Nigeria [14]. Of the seven studies conducted in Southern Nigeria, five were conducted in Southwest Nigeria (Lagos, Oyo, Ondo) [13, 15–18], two in Southsouth Nigeria [11, 12], and one in Southeast Nigeria (Enugu) [14]. Four of the studies were conducted in Lagos State [15–18] and two in Oyo State [13, 15].

Table 2 provides a brief on the sixteen studies excluded from the systematic review. Sixteen studies were excluded mainly because the study design did not answer research question [19, 20, 23, 27, 28, 29, 31, 32] or the studies was not conducted in Nigeria [23, 25, 33, 34].

Table 3 is the summary of the measured outcomes for the current study reported by the studies that met the inclusion criteria. Two studies [16, 17] had control data but one [17] had useful control data.

**Table 3.** Summary of the data provided by studies on various outcomes featured in the criteria of this review

| **Study** | **Detection rate of new cases of HIV infections** | **Acceptability (uptake) rate** | **Incidence of high-risk behaviour** | **Repeat testing rate** | **Usability rate** | **Willingness rate** | **Awareness rate** | **Incidence of social harm** |
| --- | --- | --- | --- | --- | --- | --- | --- | --- |
| Adebimpe et al (2022) [11] | NR | 306/357 (86.0%) | NR | NR | NR | NR | NR | NR |
| Agada et al (2021) [12] | 39/963 (4.04%) | 1207/5153 (23.4%) | NR | 35/871 (4.01%) | NR | NR | NR | NR |
| Babatunde et al (2022) [13] | NR | 97/155 (62.6%) | NR | NR | NR | NR | NR | NR |
| Iliyasu et al (2020) [14] | NR | 143/399 (35.8%) | NR | NR | NR | 143/399 (35.8%) | 233/399 (55.9%) | NR |
| Iwelunmor et al (2022) [15] | NR | NR | NR | NR | NR | NR | NR | NR |
| Nwoga et al (2012) [16] | 190/281 (67.6%) | NR | NR | NR | NR | NR | NR | NR |
| Ong et al (2022) [17] | NR | 180/234 (76.8%) for Oraquick vs 261/270 (96.6%) for blood testing | 2/234(0.85%) for Oraquick vs 3/270 (1.1%) for blood testing | NR | NR | NR | NR | NR |
| Tun et al (2018) [18] | 14/257 (5.6%) | NR | NR | 93/257 (36.2%) | 251/257 (97.7%) | NR | NR | NR |

### The pooled estimate of detection rate of new cases of HIV infections in Nigeria

Following confirmatory testing, the pooled detection rate of new cases of HIV infections in Nigeria using the HIV self-testing reported by the three studies [12, 16, 18] was 25.78% (95% CI: 0.90-50.66, Z=2.03, p=0.00001, I^2^: 100.0) as shown in Figure 2.

**Figure 2:**
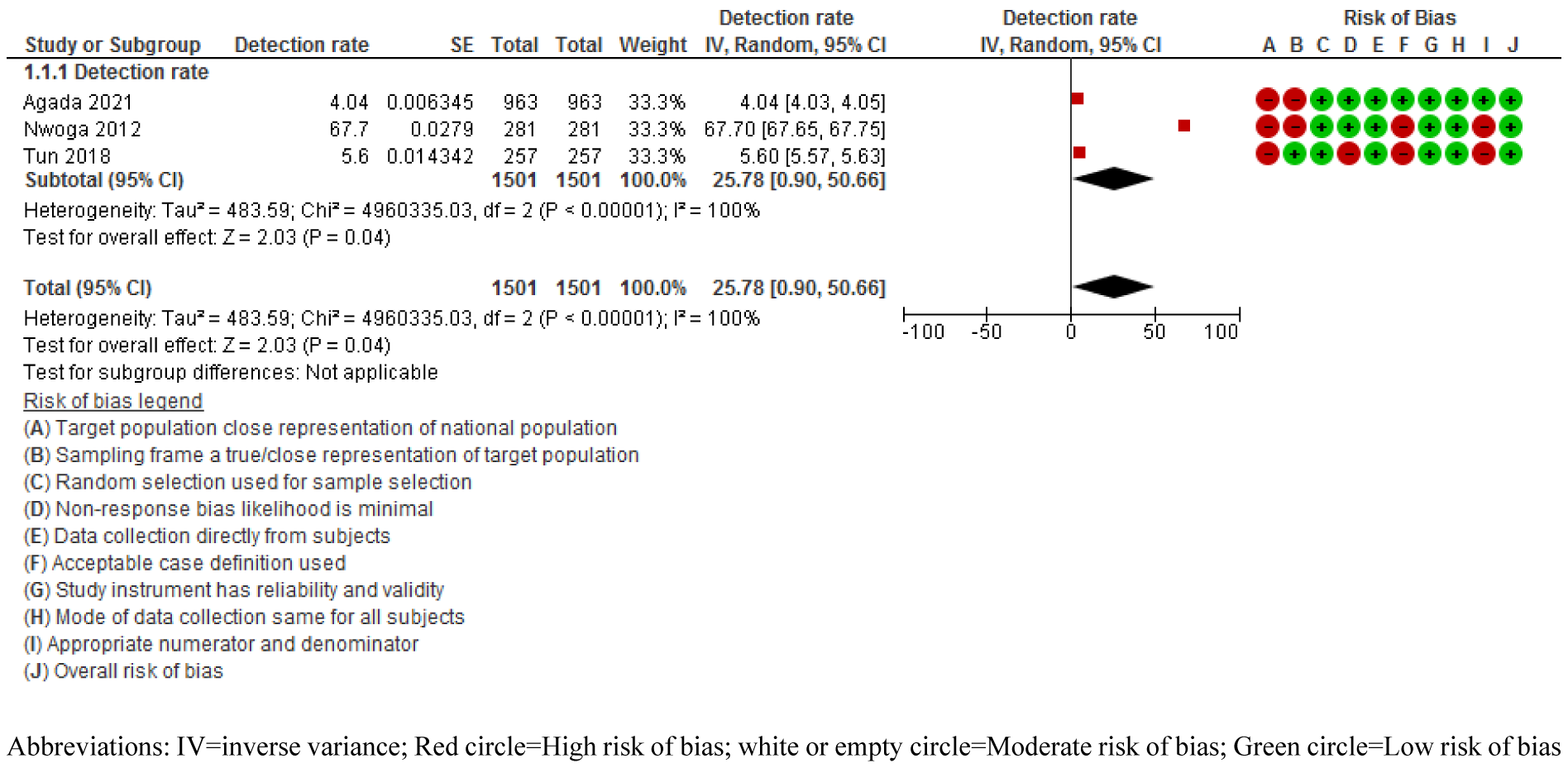
Meta-analysis showing the pooled detection rate of new cases of HIV infections in Nigeria

### The pooled estimate of acceptability (uptake) rate of HIV self-testing in Nigeria

The pooled acceptability (uptake) rate of HIV self-test in Nigeria reported by the five studies [11–14, 17] was 56.92% (95% CI: 26.54-87.30, Z=3.67, p=0.00001, I^2^: 100.0) as shown in Figure 3.

**Figure 3:**
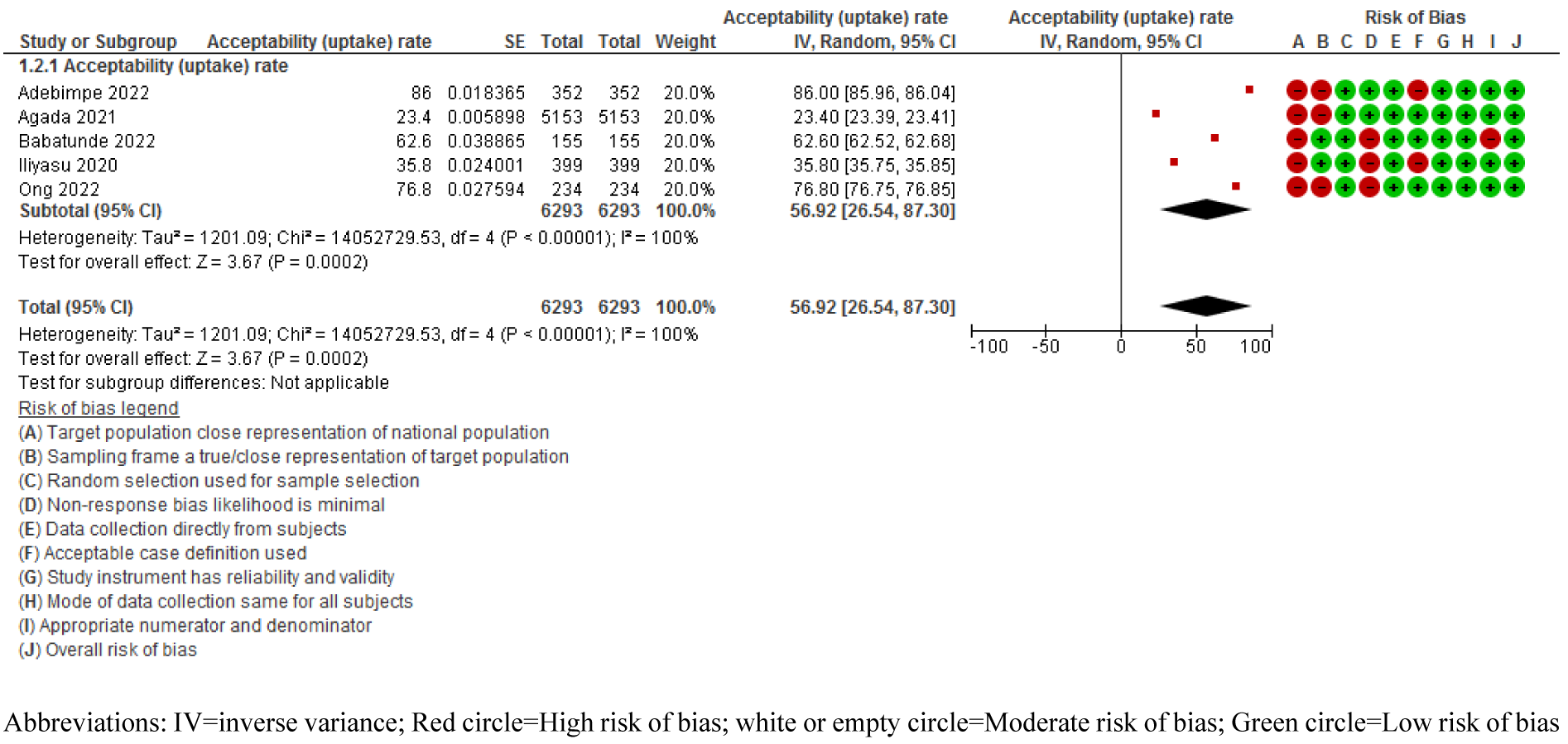
Meta-analysis showing the pooled acceptability (uptake) rate of HIV self-test in Nigeria

### The pooled estimate of usability rate of HIV self-test in Nigeria

One study reported a usability rate of HIV self-test in Nigeria to be 97.9% [18].

### The pooled estimate of repeat testing rate of HIV self-testing in Nigeria

The pooled repeat testing rate of reported by two studies [12, 18] was 20.10% (95% CI: -11.44- 51.65, Z=1.25, p=0.00001, I^2^: 100.0) as shown in Figure 4.

**Figure 4:**
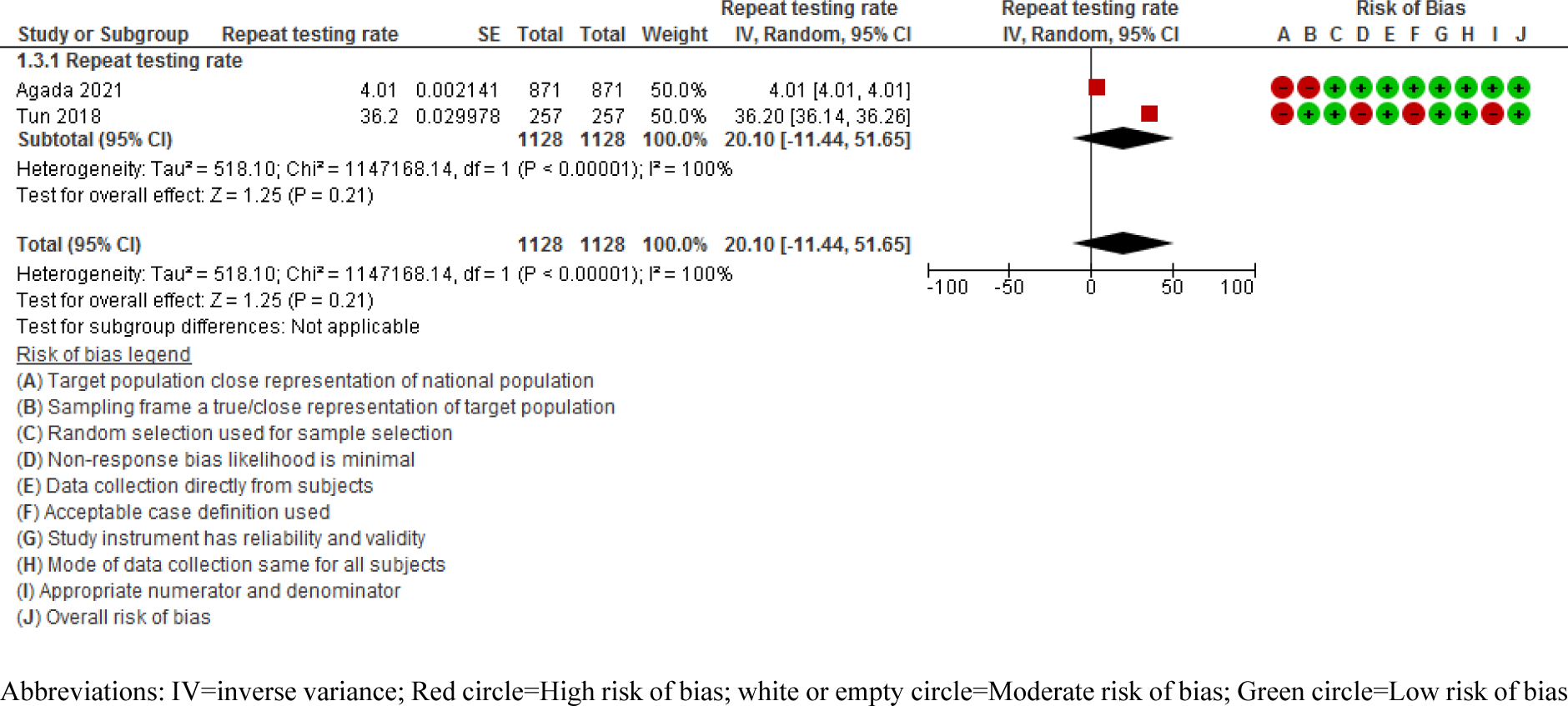
Meta-analysis showing the pooled repeat testing rate of HIV self-testing in Nigeria

### The estimate of willingness rate of HIV self-test in Nigeria

One study reported on willingness rate of HIV self-test in Nigeria at 35.8% [14].

### The estimate of awareness rate of HIV self-test in Nigeria

One study reported on awareness rate of HIV self-test in Nigeria at 55.9% [14].

### The estimate of incidence of social harm of HIV self-test in Nigeria

None of the included study reported on incidence of social harm of HIV self-testing in Nigeria.

### The estimate of incidence of high-risk behaviour of HIV self-test in Nigeria

One study reported a high-risk behaviour rate of 0.85% among the population who use HIV self-test and 1.1% for the population who used non- HIV self-test in Nigeria [17].

### Risk of Bias

Table 4 provides a summary of the risk of bias analysis. The studies included in the current systematic review and meta-analysis had low to medium risk of bias.

**Table 4:** Quality Assessment and Risk of bias scores

| Author's Name | Was the sample frame appropriate to address the target population? | Were study participants sampled in an appropriate way? | Was random selection used for sample selection? | Was the likelihood of non-response bias minimal? | Was data collection directly from subjects? | Was acceptable case definition used? | Was the study instrument of reliability and validity? | Was the mode of data collection same for all subjects? | Was there appropriate numerator and denominator? | Total Quality score | Risk of Bias |
| --- | --- | --- | --- | --- | --- | --- | --- | --- | --- | --- | --- |
| Adebimpe et al (2022) [11] | 1 | 1 | 0 | 0 | 0 | 1 | 0 | 0 | 0 | 3 | Low |
| Agada et al (2021) [12] | 1 | 1 | 0 | 0 | 0 | 0 | 0 | 0 | 0 | 2 | Low |
| Babatunde et al (2022) [13] | 1 | 0 | 0 | 1 | 0 | 0 | 0 | 0 | 1 | 3 | Low |
| Iliyasu et al (2020) [14] | 1 | 0 | 0 | 1 | 0 | 1 | 0 | 0 | 0 | 3 | Low |
| Iwelunmor et al (2022) [15] | 1 | 0 | 0 | 0 | 0 | 0 | 0 | 0 | 0 | 1 | Low |
| Nwoga et al (2012) [16] | 1 | 1 | 0 | 0 | 0 | 1 | 0 | 0 | 0 | 3 | Low |
| Ong et al (2022) [17] | 1 | 1 | 0 | 1 | 0 | 0 | 0 | 0 | 0 | 3 | Low |
| Tun et al (2018) [18] | 1 | 0 | 0 | 1 | 0 | 1 | 0 | 0 | 1 | 4 | Moderate |

### Subgroup analyses

Planned subgroup analyses based on participant age (children versus adults or adolescents and adults), sexual behaviour (sex workers versus non-sex workers), and gender (males versus females) were intended to investigate the sources of variation in the aggregated rates identified. However, there was an insufficient amount of data available to conduct additional statistical analyses.

### Sensitivity analyses

However, we performed a sensitivity analysis by excluding eligible studies individually (one by one) to explore the stability of the pooled results. The sensitivity analysis revealed that none of the inimitable studies influenced the pooled estimates.

### Publication bias assessment

Publication bias assessment could not be conducted because the number of included studies was less than 10.

### Patient and public involvement

There was no patient or public involvement in the design or execution of this systematic review and meta-analysis.

## Discussion

This systematic review and meta-analysis was needed to generate evidence on the effectiveness of ongoing efforts to reduce new HIV incidence in Nigeria through the use of innovative HIV self-testing kits like OraQuick. The outcome of the current study suggests that there was a high detection rate of new HIV cases using HIV self-testing kits in Nigeria. In addition, there was a moderate acceptability (uptake) rate but low repeat testing rate. It is important that these study findings be interpreted cautiously due to the few studies included in this analyses, significant clinical and statistical heterogeneity, and potential biases.

Furthermore, there is a paucity of published studies investigating the impact of the HIV self-testing kits on usability rate, awareness rate, willingness rate, and the incidence of high-risk behaviour. The single studies identified reported moderate rates of awareness and willingness to use HIV self-test, and a lower high-risk behaviour rate among those who use HIV self-test test than non-HIV self-test users. Furthermore, no published study addresses the incidence of social harm. One study provided control data specifically examining the impact of the HIV self-test in comparison with other HIV test kits or methods.

This is the first systematic review and meta-analysis conducted in Nigeria to determine the pooled detection rate of new HIV infection cases post-confirmatory testing, as well as the acceptability (uptake) rate of HIV self-testing and repeat testing rate. A key strength of the study is its adherence to a pre-registered protocol and the implementation of a comprehensive search strategy. In addition, most of the included studies had large sample sizes except for one that recruited fewer than 200 participants [13].

However, the review had limitations. The statistical meta-analysis was only feasible for one of the secondary outcomes due to limited published studies and available data. In addition, the available data, while of low risk of bias, had high heterogeneity. Furthermore, a wide range of study populations were included in the study though all the studies met the inclusion criteria. However, we performed a sensitivity analysis by excluding eligible studies individually (one by one) to explore the stability of the pooled results. The sensitivity analysis revealed that none of the inimitable studies influenced the pooled estimates. Despite these limitations, the study provides some important insights.

First, the high detection rate of new HIV cases using HIV self-testing kits suggest that the use of this tool can be scaled up with the potential for huge gains for the HIV response in Nigeria. Currently, the annual HIV testing coverage rate in Nigeria is high among pregnant women within the key population [35], but low among adolescents and young people [36] and low among Nigerians living in rural areas [37]. Adolescents and young people are vulnerable to HIV infection and a significant proportion of new infections occurs among adolescents and young adults in the country [38]. The ease with which HIV self-test kits can be accessed and the ability to conduct a test discretely can improve HIV testing services as these qualities help to overcome some of the barriers limiting the uptake of HIV tests [39].

Prior studies had shown that more new HIV infections are identified using HIV self-testing among key populations [40], general populations [6] and rural populations [41]. Its use increases the uptake of HIV testing when compared to standard of care [42, 43]. Uptake and use of HIV self-test increases with awareness [44]. The current study indicates that the awareness about HIV self-testing in Nigeria is only moderate, and the acceptability (uptake) of is only moderate. It is therefore plausible that HIV testing can be increased through efforts that drive awareness about HIV self-testing and the availability of HIV self-testing kits as HIV self-testing offers a pathway for populations and individuals with limited healthcare system access to take responsible actions about their HIV risk [45].

Second, a notable concern is the low frequency of repeat testing. A repeat test can be done in a clinic to confirm the result from self-testing testing to validate a positive result and facilitate appropriate linkage to prevention, treatment, and care [46]. For contexts like Nigeria where the prevalence of HIV risk behaviour is high and there is limit access to safer sex practices, repeat HIV testing is important for early detection of HIV infection and prompt access to HIV treatment [47], and for sustained effectiveness of prevention programs [48]. Though repeat testing is higher with the use of HIV self-testing than with standard HIV testing [6] the low rate of repeat testing in the current study, despite these been under experimental conditions [12, 17], is a dire call for further studies to understand the reason for the observation. Such studies can identify barriers and challenges to repeat testing and maximise the potentials that HIV self-testing has for HIV control in high burden regions like Nigeria.

Third, the observed underutilization of HIV self-testing among individuals engaged in high-risk behaviour is an interesting finding. The finding suggests that active use of HIV self-testing may be a primary preventive self-care approach for the purpose of reducing risk behaviour symptoms, or as a secondary preventive self-care approach to facilitate access to prompt treatment. These approaches reduce or even eliminates risk behaviour [49]. It may, therefore, be strategic, to market HIV self-testing as a self-care approach as this may also drive its demand and use. The self-care market has expanded into a multibillion-dollar industry driven by the connection of personal care with mental wellbeing and physical health [50].

The World Health Organization defines self-care as “the ability of individuals, families, and communities to promote health, prevent disease, maintain health, and to cope with illness and disability with or without the support of a health care provider [51]. The 2022 National HIV testing day theme promoted by the National Institute of Health was “HIV Testing is Self-Care” thereby reinforcing the self-care theme for the use of HIV self-testing [52]. Being aware of one’s HIV status empowers individuals to explore appropriate care options. There is, therefore, a critical need for research aimed at comprehending and addressing socioecological factors influencing HIV-related prevention, treatment, and care dynamics, including factors that impact access to and adoption of HIV testing and related services.

In the Nigerian context, conducting studies to develop strategies that enhance HIV testing and services becomes imperative to potentially mitigate the high rates of HIV among at-risk populations. The i-Test program for adolescents in Nigeria positions HIV self-testing as a hygiene product is an example of rebranding HIV testing for target population [14]. There are studies also exploring the utilization of HIV self-testing by adolescents in the private sector [53], the use of social network interventions for reaching hard-to-reach populations [54], the door-to-door distribution of test kit approach [55], secondary distribution of kits via peers, sexual partners, and female sex workers [56] and targeted interventions at places where at-risk populations congregate and train laypersons such as patent medicine vendors, promote rapid testing [57]. Effective combination of these context-specific strategies and interventions can improve on the use of HIV self-testing in Nigeria.

Finally, the current study identified information gaps needed to answer a few of the study questions. Information is needed on the impact of HIV self-testing tools on the incidence of social harm. A prior study had identified that the use of HIV self-testing was associated with reports of social harm in Africa [58]. Marriage breakup, verbal or physical abuse, economic hardship, blame and frustration among new concordant HIV-positive couples are some of the social harms reported associated with HIV self-testing [59]. HIV self-testing, however, increase trust and the building of stronger relationships among concordant HIV-negative couples, and women felt empowered and were assertive when offering self-test kits to their partners [59]. Social harms may be context specific [60]. For this reason, there is the need to conduct studies in Nigeria and other low-and middle-income countries to identify social harms associated with the use of HIV self-testing, the impact this may have on the acceptability (uptake) of HIV self-testing and how to mitigate this impact.

## Conclusion

This systematic review and meta-analysis provide valuable insights, and underscores the necessity for continued research, awareness campaigns, and innovative approaches to enhance the uptake and impact of HIV self-testing in Nigeria. It highlights a high detection rate of new HIV cases using HIV self-testing, signifying the potential for HIV self-testing to substantially contributions to the HIV response in Nigeria. Though the acceptability (uptake) rate was moderate, the low frequency of repeat testing raises concerns about sustaining prevention efforts. The absence of data on social harm associated with the use of HIV self-testing may also negatively affect the development of mitigation strategies for HIV self-testing uptake. Understanding the nuanced impact of HIV self-testing on individuals and relationships, coupled with targeted interventions and increased awareness, may contribute to the development of a more comprehensive and effective HIV control strategy in Nigeria. In addition, the low use of HIV self-testing among individuals engaged in high-risk behaviour is a call for strategic marketing of kits in alignment with global health perspectives.

## Authors Contributions

MOF developed the idea. MOF, GUE and FTA wrote and registered the protocol on PROSPERO. GUE, GOE and FTA undertook the literature searches. GUE and FTA screened titles, abstracts, and full text articles, extracted the data and performed the initial data analysis. GUE undertook meta-analysis. MOF, GUE, GOE and FTA wrote the first draft of the manuscript. GE read through and provided intellectual input to the document. All authors contributed to the writing of each draft of the manuscript and have approved the final submitted version. All authors critically reviewed the article, gave final approval of the version to be published, agreed on the journal to which the article has been submitted, and agreed to be accountable for all aspects of the work.

## Data Availability

All data underlying the results are available as part of the article and its accompanying data document.

## Acknowledgements

The authors would like to thank the staff of Nigerian Institute for Medical Research, Lagos, Nigeria for their enabling environment towards the conduct of this systematic review.

## Disclosure statement for publication

All authors have made substantial contributions to: conception and design of the study, or acquisition of data, or analysis and interpretation of data; drafting the article or revising it critically for important intellectual content; and final approval of the version submitted. This manuscript has not been submitted for publication in another journal.

## Ethics approval and consent to participate

Ethical approval was not applicable because it is a systematic review of primary studies.

## Consent for publication

Not applicable

---

**Appendix 1:** Search strategy for systematic review and meta-analysis on the pooled estimate of Assessing the Impact of suse of HIV self-testing on the incidence of HIV Infection in Nigeria

| Database | Example of searching strategy | Number of studies |
| --- | --- | --- |
| PubMed | – #1 "self-testing"[Title/Abstract] OR "self-test"[Title/Abstract]<br>– #2 hiv[Title/Abstract] OR aids[Title/Abstract] OR acquired human immunodeficiency syndrome[Title/Abstract] OR human immunodeficiency virus[Title/Abstract]<br>– #3 Nigeria [Title/Abstract]<br>– #4 #1 AND #2 AND #3 | 27 |
| Google Scholar | – "self-testing" OR "self-test" AND (hiv OR aids OR acquired human immunodeficiency syndrome OR human immunodeficiency virus) AND Nigeria | 990 |
| COCHRANE Library | – #1 "self-testing":ti,ab OR "self-test":ti,ab<br>– #2 hiv:ti,ab OR aids:ti,ab OR acquired human immunodeficiency syndrome:ti,ab OR human immunodeficiency virus:ti,ab<br>– #3 Nigeria:ti,ab<br>– #4 #1 AND #2 AND #3 | 0 |
| SCOPUS | – #1 "self-testing":ti,ab OR "self-test":ti,ab<br>– #2 hiv:ti,ab OR aids:ti,ab OR acquired human immunodeficiency syndrome:ti,ab OR human immunodeficiency virus:ti,ab<br>– #3 Nigeria:ti,ab<br>– #4 #1 AND #2 AND #3 | 19 |
| CINAHL | – S1 TI "self-testing" OR TI "self-test"<br>– S2 TI (hiv OR aids OR acquired human immunodeficiency syndrome OR human immunodeficiency virus)<br>– S3 TI Nigeria<br>– S4 S1 AND S2 AND S3 | 13 |
| Web of science | – #1 TI=("self-testing" OR "self-test" OR "HIV self-testing") AND (Nigeria)<br>– #2 TI=(hiv OR aids OR acquired human immunodeficiency syndrome OR human immunodeficiency virus)<br>– #3 #1 AND #2 | 33 |

## References

1. National Agency for the Control of AIDS. Spectrum Version: 6.26, 2023.

2. UNAIDS. Making the end of AIDS real: What we Mean by “Epidemic Control”. Available at: https://www.unaids.org/sites/default/files/media_asset/glion_oct2017_meeting_report_en. pdf. Accessed: 13 December 2023.

3. Oum S, Carbaugh A, and Kates J. Assessing Global HIV Targets in PEPFAR Countries: A Dashboard. Global Health Policy. Feb 10, 2021. https://www.kff.org/global-health-policy/issue-brief/assessing-global-hiv-targets-in-pepfar-countries-a-dashboard/. Accessed: 13 December 2023.

4. Onovo AA, Adeyemi A, Onime D, Kalnoky M, Kagniniwa B, Dessie M, Lee L, Parrish D, Adebobola B, Ashefor G, Ogorry O, Goldstein R, Meri H. Estimation of HIV prevalence and burden in Nigeria: a Bayesian predictive modelling study. EClinicalMedicine. 2023 Jul 20;62:102098. doi: 10.1016/j.eclinm.2023.102098.

5. Sundararajan R, Ponticiello M, Nansera D, Jeremiah K, Muyindike W. Interventions to Increase HIV Testing Uptake in Global Settings. Curr HIV/AIDS Rep. 2022;19(3):184–193. doi: 10.1007/s11904-022-00602-4.

6. Mohlabane N, Tutshana B, Peltzer K, Mwisongo A. Barriers and facilitators associated with HIV testing uptake in South African health facilities offering HIV Counselling and Testing. Health SA Gesondheid. 2016; 21: 86–95. 10.1016/j.hsag.2015.11.001.

7. Vashisht S, Jha S, Thakur N, Khaitan A, Rai S, Haldar P, et al. Comparing the Effects of Oral HIV Self-Testing With Those of Standard HIV Testing for Men Who Have Sex With Men (MSM): A Systematic Review and Meta-Analysis. Cureus. 2022 Aug 19;14(8):e28157. doi: 10.7759/cureus.28157.

8. Munn Z, Peters MDJ, Stern C, Tufanaru C, McArthur A, Aromataris E. Systematic review or scoping review? Guidance for authors when choosing between a systematic or scoping review approach. BMC Med Res Methodol. 2018 Nov 19;18(1):143. doi: 10.1186/s12874-018-0611-x.

9. Minozzi S, Cinquini M, Gianola S, Gonzalez-Lorenzo M, Banzi R. The revised Cochrane risk of bias tool for randomized trials (RoB 2) showed low interrater reliability and challenges in its application. J Clin Epidemiol. 2020 Oct;126:37–44. doi: 10.1016/j.jclinepi.2020.06.015.

10. Hoy D, Brooks P, Woolf A, Blyth F, March L, Bain C, et al. Assessing risk of bias in prevalence studies: modification of an existing tool and evidence of interrater agreement. J Clin Epidemiol. 2012 Sep;65(9):934–9. doi: 10.1016/j.jclinepi.2011.11.014.

11. Adebimpe WO, Ebikeme D, Omobuwa O, Oladejo E. How acceptable is the HIV/AIDS self-testing among women attending immunization clinics in Effurun, Southern Nigeria. Marshall J Med. 2019; 5(3): 37 DOI: 10.33470/2379-9536.1225.

12. Agada P, Ashivor J, Oyetola AB, Usang SA, Asuquo B, Nuhu T. Reaching out to the hard-to-reach populations with HIV self-testing services in South-south Nigeria. J Pre- Clin Clin Res. 2021; 15(4): 155–161. doi: 10.26444/jpccr/144699.

13. Babatunde AO, Agboola P, Babatunde Y, Ilesanmi EB, Ayodele H, Ezechi OC. Assessment of knowledge and acceptability of HIV self-testing among students of selected universities in southwest Nigeria: an online cross-sectional study. Pan Afr Med J. 2022 Oct 24;43:94. doi: 10.11604/pamj.2022.43.94.31741.

14. Iliyasu Z, Kassim RB, Iliyasu BZ, Amole TG, Nass NS, Marryshow SE, et al. Acceptability and correlates of HIV self-testing among university students in northern Nigeria. Int J STD AIDS. 2020 Aug;31(9):820–831. doi: 10.1177/0956462420920136.

15. Iwelunmor J, Ezechi O, Obiezu-Umeh C, Gbaja-Biamila T, Musa AZ, Nwaozuru U, et al. Enhancing HIV Self-Testing Among Nigerian Youth: Feasibility and Preliminary Efficacy of the 4 Youth by Youth Study Using Crowdsourced Youth-Led Strategies. AIDS Patient Care STDS. 2022 Feb;36(2):64–72. doi: 10.1089/apc.2021.0202.

16. Nwoga MC, Odukoya O, Savage KO, Effiom OA. Evaluation of accuracy of oraquick® rapid test in detecting HIV antibodies in saliva of Nigerians. Nig Dent J, 2012; 20 (2):66–69.

17. Ong JJ, Nwaozuru U, Obiezu-Umeh C, Airhihenbuwa C, Xian H, Terris-Prestholt F, et al. Designing HIV Testing and Self-Testing Services for Young People in Nigeria: A Discrete Choice Experiment. Patient. 2021 Nov;14(6):815–826. doi: 10.1007/s40271-021-00522-2.

18. Tun W, Vu L, Dirisu O, Sekoni A, Shoyemi E, Njab J, et al. Uptake of HIV self-testing and linkage to treatment among men who have sex with men (MSM) in Nigeria: A pilot programme using key opinion leaders to reach MSM. J Int AIDS Soc. 2018 Jul;21 Suppl 5(Suppl Suppl 5):e25124. doi: 10.1002/jia2.25124.

19. Adeoti AO, Desalu OO, Oluwadiya KS. Sexual practices, risk perception and HIV self-testing acceptability among long-distance truck drivers in Ekiti State, Nigeria. Nigerian Postgraduate Medical Journal. 2021 Oct 1;28(4):273–7.

20. Adepoju VA, Umebido C, Hassan Z, Adetosoye A, Olowu A, Ademola-Kay TE, et al. How efficient are HIV self-testing models? A comparison of community, facility, one-stop-shop and pharmacy retail distribution models in Nigeria. Journal of the International AIDS Society 2022; 25: 45–45.

21. Aizobu D, Wada YH, Anyanti J, Omoregie G, Adesina B, Malaba S, et al. Enablers and barriers to effective HIV self-testing in the private sector among sexually active youths in Nigeria: A qualitative study using journey map methodology. Plos one. 2023 Apr 27;18(4):e0285003.

22. Brown B, Folayan MO, Imosili A, Durueke F, Amuamuziam A. HIV self-testing in Nigeria: public opinions and perspectives. Glob Public Health. 2015; 10 (3): 354–65.

23. Dirisu O, Sekoni A, Vu L, Adebajo S, Njab J, Shoyemi E, Ogunsola S, Tun W. ‘I will welcome this one 101%, I will so embrace it’: a qualitative exploration of the feasibility and acceptability of HIV self-testing among men who have sex with men (MSM) in Lagos, Nigeria. Health Education Research. 2020 Dec;35(6):524–37.

24. Hensen B. Community-based distribution of oral HIV self-testing kits.

25. Mason S, Ezechi OC, Obiezu-Umeh C, Nwaozuru U, BeLue R, Airhihenbuwa C, et al. Understanding factors that promote uptake of HIV self-testing among young people in Nigeria: Framing youth narratives using the PEN-3 cultural model. Plos one. 2022 Jun 3;17(6):e0268945.

26. Mavhu W, Makamba M, Hatzold K, Maringwa G, Takaruza A, Mutseta M, et al. Preferences for oral-fluid-based or blood-based HIV self-testing and provider-delivered testing: an observational study among different populations in Zimbabwe. BMC Infectious Diseases. 2022 Jan;22(Suppl 1):973.

27. Negedu O, Tech B. HIV self-testing among young persons in Nigeria: Knowledge, perceptions and potential for use. InAPHA’s 2019 Annual Meeting and Expo (Nov. 2- Nov. 6) 2019 Nov 3. APHA.

28. Nwaozuru U, Gbajabiamila T, Obiezu-Umeh C, Mason S, Tahlil K, Oladele D, et al. An innovation bootcamp model to develop HIV self-testing social enterprise among young people in Nigeria: a youth participatory design approach. The Lancet Global Health. 2020 Apr 1;8:S12.

29. Nwaozuru U, Tahlil KM, Obiezu-Umeh C, Gbaja-Biamila T, Asuquo SE, Idigbe I, et al. Tailoring youth-friendly health services in Nigeria: a mixed-methods analysis of a designathon approach. Global health action. 2021 Jan 1;14(1):1985761.

30. Oladele DA, Iwelunmor J, Gbajabiamila T, Obiezu-Umeh C, Okwuzu JO, Nwaozuru U, et al. An Unstructured Supplementary Service Data System to Verify HIV Self-Testing Among Nigerian Youths: Mixed Methods Analysis of Usability and Feasibility. JMIR Formative Research. 2023 Sep 25;7:e44402.

31. Rosenberg NE, Obiezu-Umeh C, Gbaja-Biamila T, Tahlil KM, Nwaozuru U, Oladele D, et al. Strategies for enhancing uptake of HIV self-testing among Nigerian youths: a descriptive analysis of the 4YouthByYouth crowdsourcing contest. BMJ innovations. 2021 Jul;7(3):590.

32. Tahlil KM, Obiezu-Umeh C, Gbajabiamila T, Nwaozuru U, Oladele D, Musa AZ, et al. A designathon to co-create community-driven HIV self-testing services for Nigerian youth: findings from a participatory event. BMC infectious diseases. 2021 May 31;21(1):505.

33. Young SD, Daniels J, Chiu CJ, Bolan RK, Flynn RP. Acceptability of Using Electronic Vending Machines to Deliver Oral Rapid HIV Self-Testing.

34. Zhou Y, Wu D, Tang WM, Li XF, Huang SZ, Liu YW, et al. The roles of two HIV self-testing models in promoting HIV-testing among men who have sex with men. Zhonghua liu Xing Bing xue za zhi= Zhonghua Liuxingbingxue Zazhi. 2021 Feb 1;42(2):263–8.

35. Ajayi A, Awopegba O, Owolabi E, Ajala A. Coverage of HIV testing among pregnant women in Nigeria: progress, challenges and opportunities. J Public Health (Oxf). 2021 Apr 12;43(1):e77–e84. doi: 10.1093/pubmed/fdz152.

36. Ajayi AI, Awopegba OE, Adeagbo OA, Ushie BA. Low coverage of HIV testing among adolescents and young adults in Nigeria: Implication for achieving the UNAIDS first 95. PLoS One. 2020 May 19;15(5):e0233368. doi: 10.1371/journal.pone.0233368.

37. Onoja A, Onoja S, Sanni OF, Adamu I, Abiodun OP, Shaibu J. Uptake of HIV Testing: Assessing the Impact of a Community-Based Intervention in Rural Nigeria. Health Sciences and Diseases. 2020; 21 (6): 73–79.

38. UNAIDS. Global HIV & AIDS statistics—2018 fact sheet 2018. Available from: http://www.unaids.org/sites/default/files/media_asset/UNAIDS_FactSheet_en.pdf.

39. Mason S, Ezechi OC, Obiezu-Umeh C, Nwaozuru U, BeLue R, Airhihenbuwa C, Gbaja- Biamila T, Oladele D, Musa AZ, Modi K, Parker J, Uzoaru F, Engelhart A, Tucker J, Iwelunmor J. Understanding factors that promote uptake of HIV self-testing among young people in Nigeria: Framing youth narratives using the PEN-3 cultural model. PLoS One. 2022 Jun 3;17(6):e0268945.

40. Witzel TC, Eshun-Wilson I, Jamil MS, Tilouche N, Figueroa C, Johnson CC, et al. Comparing the effects of HIV self-testing to standard HIV testing for key populations: a systematic review and meta-analysis. BMC Med. 2020 Dec 3;18(1):381. doi: 10.1186/s12916-020-01835-z.

41. Nangendo J, Katahoire AR, Karamagi CA, Obeng-Amoako GO, Muwema M, Okiring J, et al. Uptake and perceptions of oral HIV self-testing delivered by village health teams among men in Central Uganda: A concurrent parallel mixed methods analysis. PLOS Glob Public Health. 2023 Jun 14;3(6):e0002019. doi: 10.1371/journal.pgph.0002019.

42. Jain S, Lowman ES, Kessler A, Harper J, Rumoro DP, Smith KY, et al. Seroprevalence study using oral rapid HIV testing in a large urban emergency department. J Emerg Med. 2012 Nov;43(5):e269–75. doi: 10.1016/j.jemermed.2012.02.021.

43. Johnson CC, Kennedy C, Fonner V, Siegfried N, Figueroa C, Dalal S, et al. Examining the effects of HIV self-testing compared to standard HIV testing services: a systematic review and meta-analysis. J Int AIDS Soc. 2017 May 15;20(1):21594. doi: 10.7448/IAS.20.1.21594.

44. Khezri M, Goldmann E, Tavakoli F. et al. Awareness and willingness to use HIV self-testing among people who inject drugs in Iran. Harm Reduct J. 2023; 20: 145. 10.1186/s12954-023-00881-z

45. Bacon A, Wang W, Lee H, Umrao S, Sinawang PD, Akin D, et al. Review of HIV Self Testing Technologies and Promising Approaches for the Next Generation. Biosensors (Basel). 2023 Feb 20;13(2):298. doi: 10.3390/bios13020298.

46. Asante AD. Scaling up HIV prevention: why routine or mandatory testing is not feasible for sub-Saharan Africa. Bulletin of the World Health Organization. 2007;85(8):644–6. 10.2471/BLT.06.037671.

47. Perkins JM, Nyakato VN, Kakuhikire B, Mbabazi PK, Perkins HW, Tsai AC, et al. Actual Versus Perceived HIV Testing Norms, and Personal HIV Testing Uptake: A Cross-Sectional, Population-Based Study in Rural Uganda. AIDS and behavior. 2018;22(2):616–28. 10.1007/s10461-017-1691-z

48. Oladele DA, Iwelunmor J, Gbajabiamila T, Obiezu-Umeh C, Okwuzu JO, Nwaozuru U, et al. An Unstructured Supplementary Service Data System to Verify HIV Self-Testing Among Nigerian Youths: Mixed Methods Analysis of Usability and Feasibility. JMIR Form Res. 2023 Sep 25;7:e44402. doi: 10.2196/44402.

49. Petriková F, Lichner V, Žiaková E. Risk Behavior Versus Self-Care in Adolescent. European Journal of Social Science Education and Research. 2020; 7(3): 62–73.

50. Kerfoot. What is driving driving the expanding personal care market for ‘self care’. ND. Available at: https://www.kerfootgroup.co.uk/what-is-driving-the-expanding-personal-care-market-for-self-care. Accessed: 13 December 2023.

51. World Health Organization. July 2021. WHO Guideline on Self-Care Interventions for Health and Well-Being. Available at https://www.who.int/publications/i/item/9789240052192.

52. National Institute of Health. National HIV Testing Day—Practice Self-Care with an HIV Test. October 18, 2022. Available at: https://www.oar.nih.gov/about/directors-corner/national-hiv-testing-day-practice-self-care-hiv-test#:~:text=The%20NIH%20OAR%20encourages%20universal,and%20protect%20their%20loved%20ones. Accessed: 13 December, 2023.

53. Aizobu D, Wada YH, Anyanti J, Omoregie G, Adesina B, Malaba S, et al. Enablers and barriers to effective HIV self-testing in the private sector among sexually active youths in Nigeria: A qualitative study using journey map methodology. PLoS ONE, 2023; 18(4): e0285003. 10.1371/journal.pone.0285003

54. Sheira LA, Kwena ZA, Charlebois ED, Agot K, Ayieko B, Gandhi M, et al. Testing a social network approach to promote HIV self-testing and linkage to care among fishermen at Lake Victoria: study protocol for the Owete cluster randomized controlled trial. Trials. 2022 Jun 6;23(1):463. doi: 10.1186/s13063-022-06409-3.

55. Indravudh PP, Fielding K, Chilongosi R, Nzawa R, Neuman M, Kumwenda MK, et al. Effect of door-to-door distribution of HIV self-testing kits on HIV testing and antiretroviral therapy initiation: a cluster randomised trial in Malawi. BMJ Glob Health. 2021 Jul;6(Suppl 4):e004269. doi: 10.1136/bmjgh-2020-004269

56. Hamilton A, Thompson N, Choko AT, Hlongwa M, Jolly P, Korte JE, et al. HIV Self-Testing Uptake and Intervention Strategies Among Men in Sub-Saharan Africa: A Systematic Review. Front Public Health. 2021 Feb 19;9:594298. doi: 10.3389/fpubh.2021.594298.

57. Chiu C, Hunter LA, McCoy SI, Mfaume R, Njau P, Liu JX. Sales and pricing decisions for HIV self-test kits among local drug shops in Tanzania: a prospective cohort study. BMC Health Serv Res. 2021 May 6;21(1):434. doi: 10.1186/s12913-021-06432-1.

58. Njau B, Damian DJ, Abdullahi L, Boulle A, Mathews C. The effects of HIV self-testing on the uptake of HIV testing, linkage to antiretroviral treatment and social harms among adults in Africa: A systematic review and meta-analysis. PLoS One. 2021 Jan 27;16(1):e0245498. doi: 10.1371/journal.pone.0245498.

59. Kumwenda MK, Johnson CC, Choko AT, Lora W, Sibande W, Sakala D, et al. Exploring social harms during distribution of HIV self-testing kits using mixed-methods approaches in Malawi. J Int AIDS Soc. 2019 Mar;22 Suppl 1(Suppl Suppl 1):e25251. doi: 10.1002/jia2.25251.

60. Sanaullah. Context-specificity of violence: Physical, psychological, and social dimensions of harm during the Taliban’s insurgency (2007–2009) in Pakistan. Violence: An International Journal. 2021; 2(1): 43–64. 10.1177/2633002421994818.

